# Adoption of AI-Powered Chatbots with Large Language Models by Pathologists

**DOI:** 10.1101/2024.04.05.24305405

**Authors:** Andrey Bychkov, Thiyaphat Laohawetwanit, Daniel Gomes Pinto

## Abstract

**Aims:** The study aimed to investigate the adoption and perception of artificial intelligence (AI) chatbots, particularly those powered by large language models (LLMs), among pathologists worldwide. It explored the extent of their engagement with these technologies, identifying potential impacts on their professional practices.

**Methods:** A cross-sectional survey was conducted, gathering data from pathologists on their usage and views concerning AI chatbots powered by LLMs. The survey, distributed globally via various digital platforms, included both quantitative and qualitative questions. Statistical analyses were performed to delineate patterns in the adoption and perspectives on these AI tools among the respondents.

**Results:** Of 215 respondents, 100 (46.5%) reported using LLMs, particularly ChatGPT, for professional purposes, predominantly for information retrieval, proofreading, and academic writing, highlighting a significant time-saving benefit. The adoption varied across demographics, with younger, male pathologists showing higher usage rates. While the technology was mainly utilized for drafting academic materials and programming tasks, users expressed concerns about information accuracy, privacy, and the need for regulatory approval. Despite recognizing occasional inaccuracies, respondents saw potential in advanced AI features, particularly in image analysis and speech-to-text functions.

**Conclusions:** The survey underscored pathologists’ cautious yet growing interest in leveraging LLMs to enhance information accessibility, efficiency, and medical education. While the potential benefits are recognized, significant apprehensions about the reliability, ethics, and security associated with these AI tools underscore the need for comprehensive regulation and standardized practices to ensure their responsible use in the medical field.

## INTRODUCTION

Artificial intelligence (AI) has recently gained public attention due to advanced deep-learning models that can create content with minimal human involvement, from art to term papers. This has sparked discussions about AI’s current and potential roles in various aspects of life. Among the many fields where AI can be applied, medicine, in particular, holds immense promise and significant challenges (1). Cutting-edge technologies like digital imaging, sophisticated AI algorithms, and computer-aided diagnostics have driven a notable transformation in diagnostic pathology. These innovations are crucial in advancing computational histopathology and AI-powered diagnostics, especially in precision medicine for cancer (2).

A chatbot is computer software that utilizes AI and natural-language processing to interpret queries and automate responses, emulating human interaction. In order to utilize a chatbot, an individual initiates a “session” by inputting a query, also known as a “prompt,” using simple and natural language. The user is generally a human being. The chatbot quickly generates a relevant natural-language “response” to the prompt, typically within 1 second. The ongoing interaction of ideas and responses persists throughout the session, resulting in a conversational dynamic akin to a dialogue between two individuals (3). Chatbot technology has become ubiquitous, being used in several domains, such as customer service and personal virtual assistants. Modern computers possess immense computational capacity, enabling large language models (LLMs) to contain hundreds of billions of parameters. These parameters can be utilized to generate fresh textual content. The capacity to utilize a vast quantity of accessible (Internet) data for training the network allows language models to expand their capabilities continuously (4).

Several AI chatbots, including ChatGPT and BingAI, are widely available and actively developed by AI research and development companies. These tools were not specifically built for medical note analysis or image recognition tasks. Instead, they were designed to possess versatile cognitive abilities to assist users in accomplishing various tasks. A prompt may be an inquiry or a directive to carry out a particular action. In addition, prompts are not limited to being sentences in English. They can be written in several human languages and may include data inputs such as spreadsheets, technical specifications, research papers, and mathematical equations (3). Despite recent advancements, there is a lack of data on how pathologists currently apply these technologies to their professional lives. This multinational survey aims to evaluate global pathologists’ engagement with AI-driven chatbots utilizing LLMs.

## METHODS

### Survey design and distribution

This study employed a cross-sectional survey design to gather data from pathologists regarding their use and perception of AI equipped with LLMs such as ChatGPT and Gemini. The survey was created using Google Forms due to its accessibility and ease of use. The questionnaire comprised closed-ended and open-ended questions designed to elicit detailed responses about the participants’ experiences with LLMs in their professional practice.

The survey was disseminated globally through various channels over eight months, from August 2023 to March 2024. These channels included social media platforms (LinkedIn, X, and Facebook), direct email invitations to the professional network of the authors, and announcements in pathology-related online forums and newsletters. The aim was to reach a diverse and representative sample of pathologists and pathology trainees.

### Participant selection

Participants were eligible for the study if they were practicing pathologists or pathology trainees. There were no restrictions based on geography, age, gender, or experience level. The survey was available in English, and participants were informed that their responses would be used for research purposes. Consent was obtained electronically before participation in the survey.

### Data collection and analysis

Data collection occurred over eight months, allowing adequate time for many participants to respond. Upon closure of the survey, responses were exported from Google Forms for analysis. The data was analyzed using statistical software. Descriptive statistics, such as frequencies and percentages, were used to summarize the participants’ demographic characteristics and responses to closed-ended questions. Chi-square tests calculated the difference between each subgroup. Statistical significance was established at p-values of less than 0.05. Open-ended responses were analyzed qualitatively to identify common themes and perspectives regarding using LLMs in pathology.

## RESULTS

### Respondent characteristics

A total of 215 individuals participated in the study, as shown in Table 1. Among these, 100 pathologists (46.5%) employed LLMs or chatbots in their professional activities, whereas 115 participants (53.5%) did not. The latter group featured a more females and individuals aged between 41 and 55 years compared to the group of users, with this difference being statistically significant (*P*=0.03).

**Table 1:**
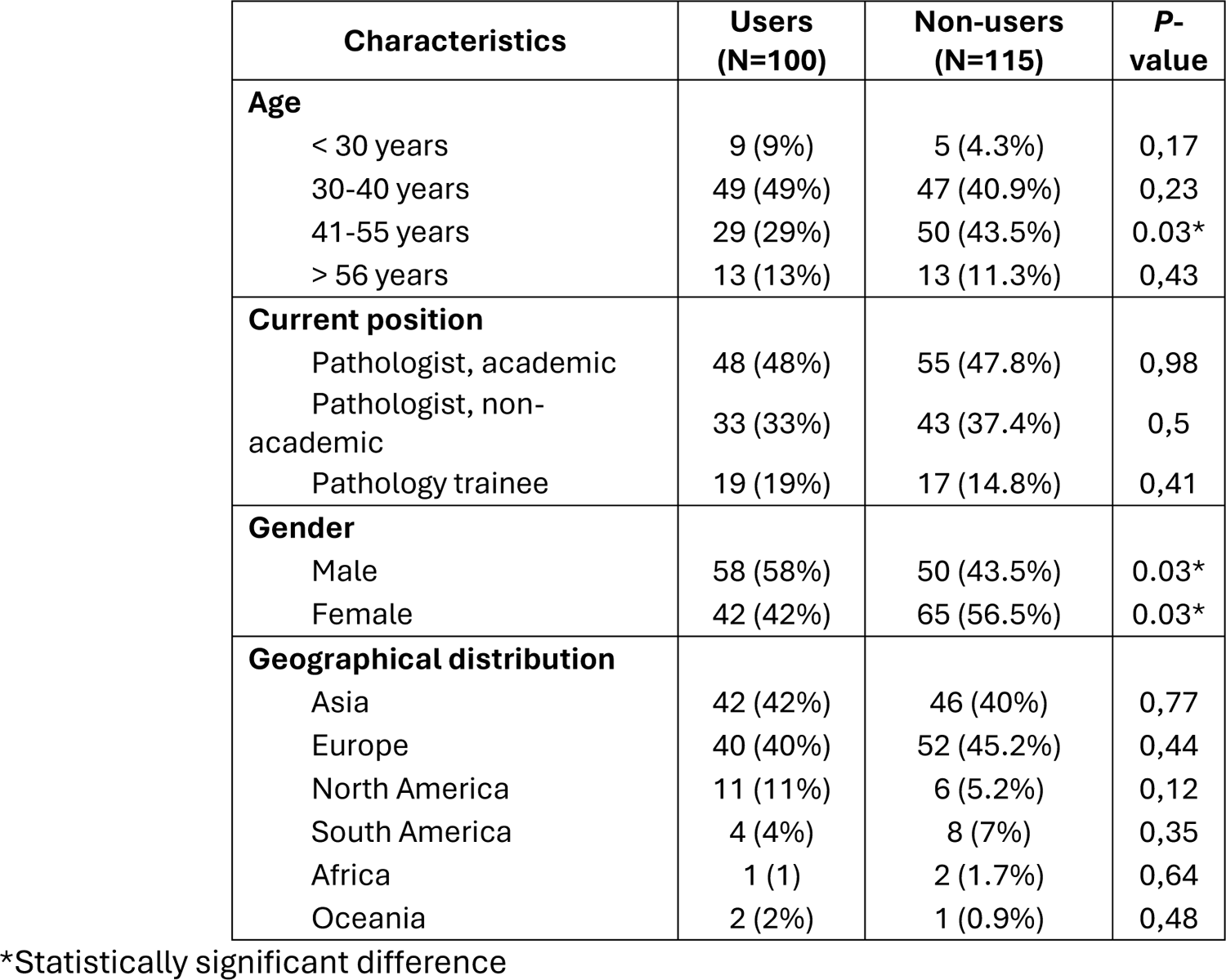
Respondent characteristics.

The user group exhibited a significantly greater understanding of coding skills, generative AI, and LLMs than the non-user group (*P*<0.01). In particular, academic pathologists demonstrated the highest level of understanding in digital pathology and LLMs (*P*=0.02). Conversely, pathology trainees who constituted about 20% of the users cohort reported lower levels of understanding, particularly in digital pathology and LLMs.

### Chatbot usage among user-group

Table 2 presents data regarding the utilization of chatbots by 100 pathologists who use these application in their work. The majority, 76 users, favored the cost-free ChatGPT (GPT-3.5), whereas a smaller cohort of 12 users opted for the subscription-based GPT-4. Additionally, 37 participants were familiar with and used several different chatbots. The frequency of chatbot usage was diverse: 40 users engaged with chatbots 5-10 times per week, 24 interacted with them several times a day, and 36 used them less frequently. Regarding access methods, 52 participants used computers, while 39 utilized both computers and mobile applications. The predominant applications of chatbots were for information retrieval and assistance with proofreading or translation (Table 1). Furthermore, chatbots were notably employed in the academic domain, particularly for initial drafting and editing of the manuscripts. More than half of the respondents reported saving 1 to 5 working hours per week as a result of using the chatbot applications.

**Table 2:** Use of LLMs among user group.

| LLMs usage | Respondents<br>(N=100) |
| --- | --- |
| **1. LLMs used** |  |
| ChatGPT (free/GPT-3.5) | 76 |
| ChatGPT (paid/GPT-4) | 34 |
| Bing AI | 22 |
| Google Bard | 13 |
| Several chatbots | 37 |
| **2. Frequency** |  |
| 5-10 times per week | 40 |
| Rarely | 36 |
| Several times a day | 24 |
| **3. Accessing LLMs** |  |
| Only personal computers | 52 |
| Both personal computers and mobile applications | 39 |
| Only mobile applications | 9 |
| **4. Use cases** |  |
| Information search (i.e., googling) | 66 |
| Correspondence (i.e., proofreading, translation) | 65 |
| Academic work: proofreading and polishing manuscripts | 58 |
| Academic work: drafting | 53 |
| Creation of educational content (i.e., MCQs, presentations, cases) | 43 |
| Coding | 23 |
| LLM-based research | 20 |
| Writing pathological reports | 16 |
| **5. Use of LLMS for creation and promotion of academic work** |  |
| Take-away summaries | 49 |
| Abstracts | 47 |
| Titles | 36 |
| Images, icons, and other visuals | 27 |
| Social media posts | 24 |
| **6. Use of LLMs to help with the drafting of academic manuscripts** |  |
| Not used LLMs | 51 |
| Original studies | 32 |
| Shorts pieces (e.g., editorials, perspectives, case reports) | 28 |
| Reviews (e.g., narrative, systematic, scoping, etc.) | 24 |
| **7. Use of LLMs in different sections of academic manuscripts** |  |
| Introduction | 24 |
| Abstract | 23 |
| Discussion | 21 |
| Conclusion | 18 |
| Materials and methods | 13 |
| Results | 13 |
| Figures | 5 |
| **8. Use of GPT-4 as an aid in programming tasks** |  |
| Writing simple command-line programs/scripts | 20 |
| Coding tutorial | 14 |
| Code clean-up | 11 |
| Simple GUI-based applications | 9 |
| Machine learning algorithms | 9 |
| Complex applications | 6 |
| Code commenting | 6 |
| **9. Performance in different languages compared with English** |  |
| No experience | 36 |
| Same | 28 |
| Worse | 26 |
| Better | 6 |
| **10. Quality of translation compared with mainstream applications** |  |
| Better | 42 |
| Same | 25 |
| No experience | 22 |
| Worse | 6 |

A subgroup analysis focusing on pathologists who applied LLMs in their academic work revealed that the primary application was in writing the introduction and abstract sections, with 24 and 23 participants utilizing LLMs for these parts, respectively. Approximately 30 participants indicated that LLMs contributed 1-25% of content in some of their manuscripts. At the same time, 37 pathologists familiar with ChatGPT reported no usage of LLMs in their published works.

In a subgroup of 24 participants who employed GPT-4 for programming-related tasks, LLMs were predominantly utilized for developing command-line programs and scripts, as reported by 20 users, followed by creating coding tutorials (14 users) and performing code clean-up (11 users). GPT-4 was deemed moderately reliable for these coding tasks, receiving an average reliability rating of 3.4 out of 5. In terms of efficiency, it was highly valued for its time-saving capabilities, with an average rating of 4.1 out of 5. Additionally, among 38 respondents who used plugins, 20 users favored web-search plugins, and 17 users chose PDF readers. The participants expressed a moderately positive stance on the necessity of regulatory approval for using LLMs in medical diagnostics, with an average score of 3.36 out of 5.

### Perception of LLMs performance

Chatbots sometimes provided incorrect general domain information, as reflected by an average score of 2.82 on a five-point scale. However, LLMs were generally perceived to possess moderate to good overall knowledge, with an average score of 3.38. Concerning pathology-specific knowledge, chatbots were considered moderately proficient, receiving an average score of 2.72. Most respondents (n = 54) reported a similar performance of chatbots in languages other than English. Additionally, when the translation capabilities of chatbots were compared to those of mainstream translation applications (e.g. Google Translate, DeepL, etc.), 42 respondents deemed the translations provided by chatbots superior.

### Limitations and concerns about LLMs

Figure 1 summarizes the limitations and concerns regarding LLMs as reported by users and non-users, respectively. Among the 100 users, the prevalent concern was providing outdated information, which is a feature of GPT-3.5, cited by 55%. Other reported limitations were issues with context capability (44%) and concerns regarding privacy and trustworthiness of responses (41% for each). In contrast, the 115 non-users primarily emphasized medicolegal and ethics issues (47.8%, 55 responses) and the risk of data misinformation (46.1%, 53 responses) when potentially employing LLMs in professional contexts.

**Figure 1:**
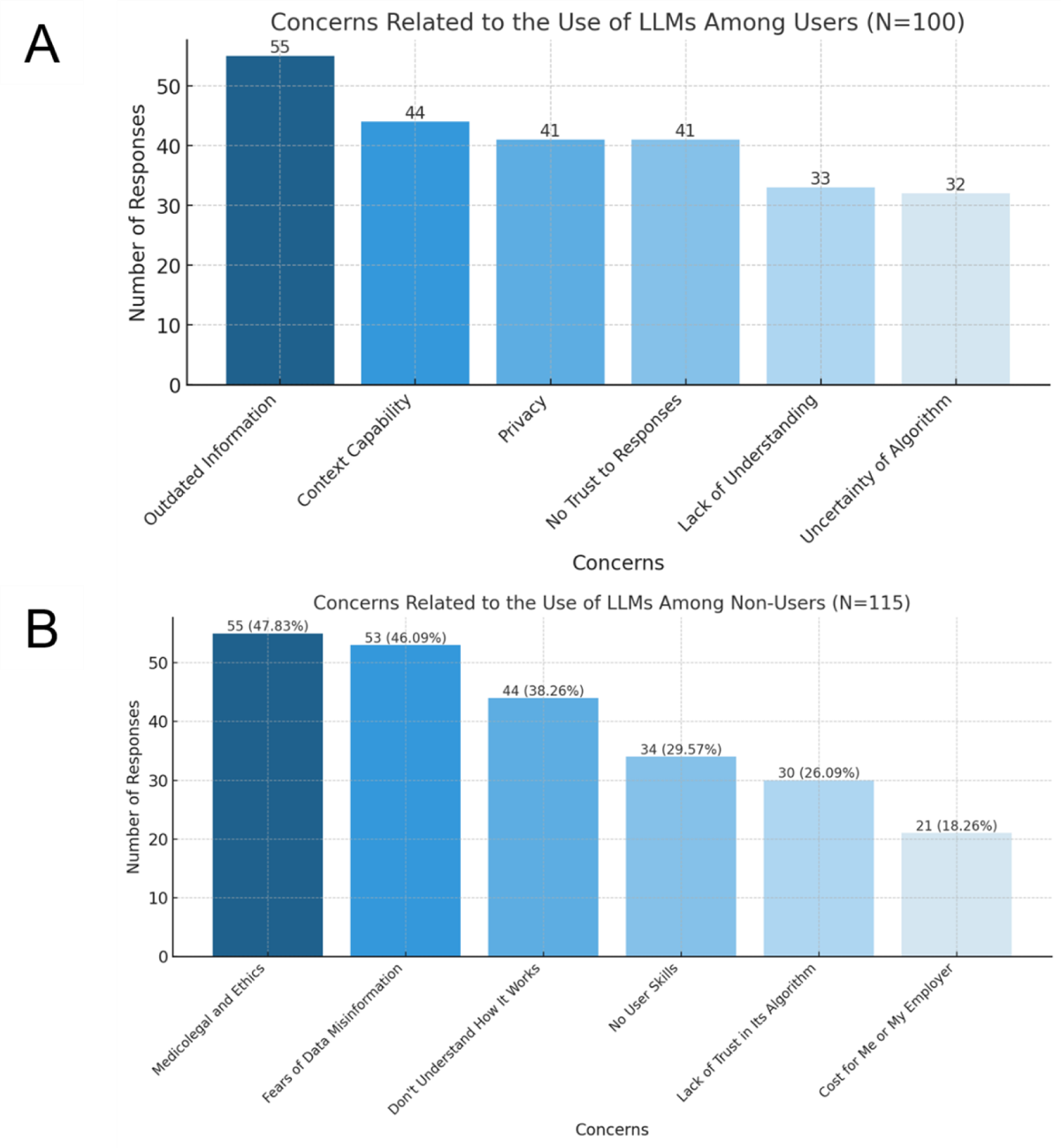
Concerns related to the use of large language models (LLMs) among (A) user group and (B) non-user group.

### Improvement of LLMs

Most respondents, 81 out of 100 (81%), indicated that they manually refined their prompts by posing additional questions. Regarding the most sought-after features in chatbots or LLMs, 61 respondents highlighted the importance of image analysis capabilities (Figure 2A). The demand for speech-to-text functionality, internet connectivity, and the ability to produce media outputs was also significant, with each receiving 52 responses. Among the 115 non-users, the primary enhancements desired for chatbots were the reliability of the output data (50.4%, 58 responses) and improved context capability (49.6%, 57 responses), as illustrated in Figure 2B.

**Figure 2:**
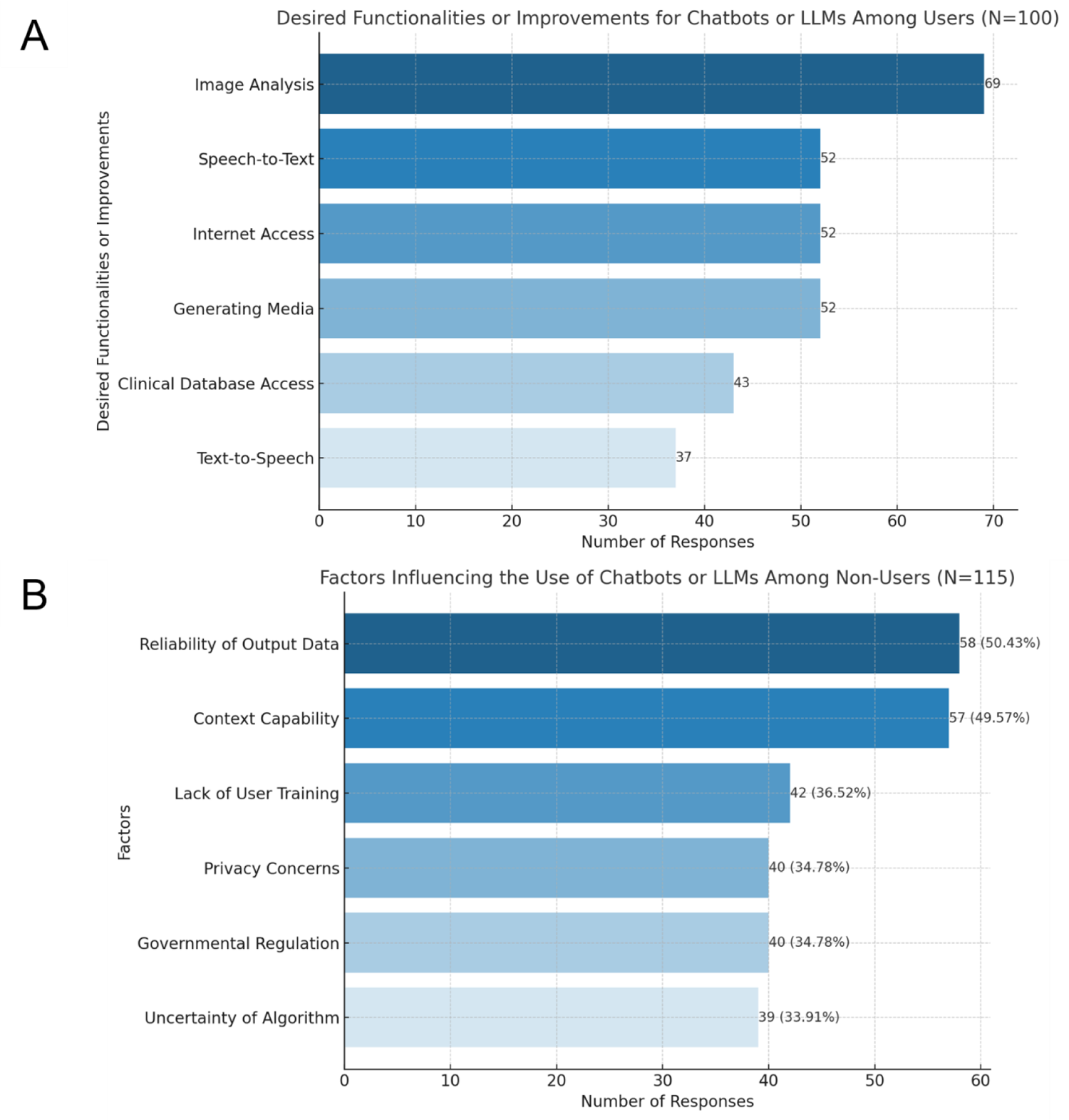
Expectation to chatbots or large language models (LLMs) among (A) user group and (B) non-user group.

### Ethical consideration

In the survey, 65 respondents refrained from sharing patient information with LLMs. Among the 32 participants who shared information, all ensured medical record numbers were anonymized, 31 anonymized demographic data, and 13 anonymized medical history. Concerning submitting unanonymized data, 70 respondents were amenable to sharing medical history, and 18 didn’t mind to share demographic data. However, no respondents advocated the sharing of medical record numbers without anonymization.

### Additional comments

Additional comments are displayed in Table 3. In summary, integrating AI in healthcare, primarily through tools such as ChatGPT, poses reliability, privacy, and practicality challenges. Although AI proves helpful for general tasks, its limitations in specialized medical settings and concerns regarding the privacy and safety of patient data highlight the necessity for prudent application and specialized training for healthcare practitioners.

**Table 3:**
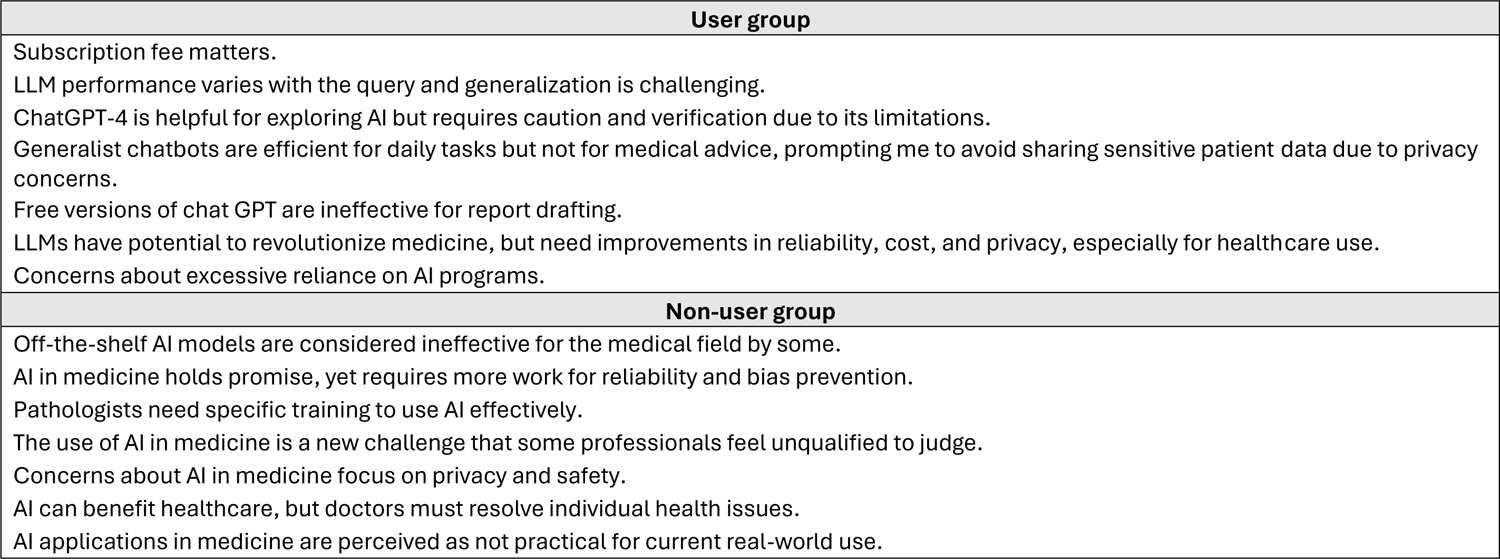
Summary of additional comments.

## DISCUSSION

The survey results indicate a varied level of engagement with LLMs and chatbots among pathologists, as approximately half of them incorporate these technologies into their professional routines. The most commonly used chatbot was ChatGPT. The adoption was notably higher among academic pathologists and younger professionals, who exhibited a substantial understanding and utilization of these tools. They predominantly employed LLMs and chatbots for tasks such as information retrieval, proofreading, translation, and academic writing, highlighting the advantages of time efficiency. Nonetheless, there were prevailing concerns among users and non-users about the risks of outdated information, the capabilities of these tools in understanding context, privacy issues, and the ethical implications, particularly concerning handling patient data. Despite these reservations, there was evident interest in advanced features like image analysis and speech-to-text functions. The general sentiment was cautious optimism, acknowledging the potential benefits of LLMs while emphasizing the necessity for enhancements in reliability, context comprehension, and ethical standards to facilitate their safe and effective integration into healthcare practices.

Our study is the first international survey to explore how pathologists and pathology trainees use AI chatbots. Consistent with earlier studies, our research indicates that nearly half of the respondents have experimented with these AI programs (5, 6). The medical profession broadly anticipates incorporating AI into healthcare, acknowledging its prospective advantages while considering the inherent limitations and challenges (7, 8). According to a survey at a medical school, approximately 33% of the faculty used ChatGPT, mainly for creating multiple-choice questions (9). While medical students perceive ChatGPT positively for its potential for treatment guidance and educational purposes, established physicians exhibit a more reserved stance toward its application (10). A survey involving 200 researchers, mainly from the medical field, revealed that while many know ChatGPT, only 11.5% have used it in their research, mainly for rephrasing text and sourcing references (11).

The utilization of LLMs by survey respondents predominantly encompassed tasks such as information retrieval, text editing, and language translation. Nearly half of the participants in the user group reported using LLMs to assist in drafting academic manuscripts, a figure consistent with prior research (6). A significant concern is using AI tools in scientific writing and the potential for cheating during training assessments (6, 12). Notably, the current study found that nearly 70% of participants who used these tools for academic writing did not disclose their use of generative AI in their publications. Several studies underscore the substantial challenge of differentiating between AI-generated and human-written content. A study evaluating AI text detectors’ ability to identify human-written text, using thousands of abstracts from scientific journals between 1980 and 2023, found that up to 8% of genuine human-generated abstracts were incorrectly marked as AI-generated (13). Additionally, studies revealed that human reviewers and AI detectors struggled to identify AI-generated abstracts accurately. Human accuracy ranged from 31.7% to 76.2%, and AI detectors were not only inefficient but could be easily bypassed with simple modifications (14, 15). Despite utilizing AI tools like ChatGPT in scientific publishing, they are not recognized as coauthors due to their inability to be accountable or possess legal capabilities. Authors are advised to clearly state their use of AI in their work, verify the accuracy of the information presented, and detail the steps they have taken to avoid plagiarism and errors in scientific publications that include AI (16).

Image analysis was the most desired functionality, as stated by approximately 70% of participants in the present study. Although studies using ChatGPT in the examination setting have shown promising results (17, 18), a few others using this AI tool for image analysis reveal mixed results. A study evaluating the ability of ChatGPT-4 in detecting and classifying colorectal adenomas from histopathological images showed that ChatGPT-4 had a high sensitivity but low specificity in identifying adenomas. The accuracy in classifying different types of polyps varied, with a generally low diagnostic consistency (19). Another study showed that ChatGPT-4 could accurately identify the presence of fatty liver disease and grades of fibrosis (20). The other found that ChatGPT-4 could identify normal blood cells with 88% accuracy and abnormal cells with 54% accuracy, slightly outperforming the manual method’s 49.5% accuracy for abnormal cells (21). Regarding these mixed results, there is still a gap between expectation and reality. Despite its potential as an auxiliary tool in image analysis, ChatGPT-4 cannot yet replace professional medical judgment.

ChatGPT is recognized for its advanced natural language comprehension, widespread availability, and adaptability in various fields such as healthcare, education, and creative arts. It provides innovative and time-efficient solutions, benefiting from its continuous learning and contextual understanding capabilities (22). However, the present study highlighted prevalent concerns associated with the use of LLMs, including the dissemination of outdated information, risks of data inaccuracies, and medicolegal challenges. These issues align with inherent difficulties faced by LLMs, such as maintaining coherence over extended interactions and the potential generation of biased or misleading content, a byproduct of its training datasets. Additionally, ethical considerations, security vulnerabilities, and privacy concerns are significant challenges that must be addressed (23).

The survey indicates a future for LLMs in pathology, emphasizing the need for education and addressing ethical and privacy concerns, particularly with patient data. It points to LLMs’ potential in tasks like information searches and academic writing, alongside a demand for features like image analysis tailored to pathology’s needs. Despite the optimism, the technology’s limitations are recognized, such as challenges in integrating digital slides and addressing biases (24). A study showing ChatGPT’s value as a consultative tool, though not a substitute for professional judgment, indicates its potential with further AI advancements (25). Concurrently, the sector faces challenges like understanding disparities, ethical concerns, and the accuracy of AI, underscoring the need for regulatory frameworks and continuous improvement to ensure LLMs’ responsible, effective, and ethical use in pathology, which involves collaboration among stakeholders to enhance patient care (26, 27).

This study has several limitations. Firstly, the online survey’s focus might bias the results toward more technologically adept pathologists, potentially affecting reliability. Additionally, the cross-sectional design limits the ability to capture the evolving usage of LLMs in pathology. Moreover, the swift progress in language model technology might soon render the study’s findings outdated. The exclusive use of English in the survey could also overlook cultural diversity. These concerns emphasize the importance of cautious result interpretation and advocate for future research to encompass broader, more diverse, and longitudinal studies to gain a comprehensive understanding of chatbots and LLMs in pathology.

In conclusion, this multinational survey has shed light on the current adoption rates, perceptions, and potential future applications of LLMs like ChatGPT among pathologists. The key findings reveal a cautious but growing interest in integrating LLMs into pathology practice, highlighting the potential for these tools to enhance information accessibility, improve efficiency, and support medical education and research. However, the study also underscores significant concerns regarding LLMs’ reliability, ethics, and privacy, emphasizing the need for regulation, particularly when used in clinical practice.

## Data Availability

All data produced in the present study are available upon reasonable request to the authors.

## Acknowledgements

The authors would like to thank Casey P. Schukow, DO (Corewell Health’s Beaumont Hospital, Royal Oak, MI) for his help with the survey creation and recruitment of respondents.

## Ethics approval statement

This study received ethical approval from the human research ethics committee of Thammasat University (Medicine), with the approval number being 66/2024.

## Contributorship statement

AB: Conception, design of the study, acquisition and analysis of data, reviewing the manuscript and figures; TL: Acquisition and analysis of data, drafting the manuscript and figures; DP: design of the study, acquisition and analysis of data, reviewing the manuscript and figures.

